# Obesity in children is associated with increased dengue virus binding, but not neutralizing, antibody responses following primary infection

**DOI:** 10.64898/2026.08.14.26360483

**Authors:** Reinaldo Mercado-Hernandez, Sandra Bos, Angel Balmaseda, Guillermina Kuan, Eva Harris

## Abstract

**Background:** Obesity has been associated with higher risk of dengue virus (DENV) infection and disease, yet its influence on antibody responses to DENV remains undefined.

**Methods:** We evaluated whether nutritional status—based on BMI z-score (BMIz)—or blood markers of body fat—leptin and adiponectin—are associated with binding and/or neutralizing antibody responses to DENV in 85 children in the Nicaraguan Pediatric Dengue Cohort Study who experienced a primary DENV infection in 2019. Associations were estimated using linear models adjusting for age, sex, and DENV infection outcome.

**Results:** Compared to children with normal weight, those with obesity had higher quantities of DENV binding antibodies (fold-change [FC] 1.89, 95% confidence interval [CI] 1.02, 3.48) but no difference in neutralizing antibodies. Likewise, leptin concentration was associated with higher quantities of binding antibodies (FC 1.22, 95%CI 1.09, 1.37), while adiponectin was associated with lower quantities (FC 0.79, 95%CI 0.67, 0.94), and neither was associated with neutralizing antibodies. Lower neutralizing efficiency (neutralizing/binding antibodies) was observed in children with obesity (FC 0.67, 95%CI 0.48, 0.93).

**Conclusions:** Our results indicate that obesity is associated with higher antibody quantity (binding) but not higher quality (neutralization) post-primary DENV infection—implying that antibodies generated by children with obesity have lower neutralization efficiency, requiring greater quantities to reach similar levels of neutralization than children with normal weight. Further, the agreement among the three models using distinct proxies of body fat—BMIz, leptin, and adiponectin—demonstrates that adipokines are useful in supplementing BMIz analysis or as independent predictors of immune responses.

**Key points:** Children with obesity in a Nicaraguan cohort had a higher magnitude of binding, but not neutralizing, antibodies to dengue virus post-primary infection, resulting in lower neutralizing efficiency. Leptin and adiponectin were effective blood-based proxies for body fat in immune studies.

## INTRODUCTION

The four dengue viruses (DENV1-4), the etiological agents of the most prevalent human mosquito-borne viral disease, infect over 100 million people annually, resulting in an estimated 50 million dengue cases [1]. Dengue is a febrile disease whose global incidence is on the rise [1]. It can progress to Dengue Hemorrhagic Fever/Dengue Shock Syndrome or Severe Dengue, which, without appropriate supportive care, can be fatal [2,3]. The development of a dengue vaccine has been challenging due to the risk of non-neutralizing DENV-binding antibodies enhancing the risk of disease in a subsequent DENV infection, via a process known as antibody-dependent enhancement (ADE) [4–6].

Recently, non-immunological host factors, specifically obesity, have been associated with higher risk of DENV infection, development of dengue disease, and progression to severe dengue [7–9]. These findings can be explained by the observed association between obesity and low-grade chronic inflammation [10,11]. Leptin, a hormone primarily produced by adipose tissue whose serum concentration positively correlates with fat mass, has been shown to impair human B-cell function [12–14]. Adiponectin is a hormone primarily synthesized by adipocytes, which, in contrast to leptin, is associated with lower body fat [15]. Given the substantial rise in the global prevalence of obesity in children and adolescents, it is urgent to evaluate the role of nutritional status, such as obesity, on pediatric infectious diseases and immune responses [16,17].

Here, we investigated whether nutritional status influences the antibody response to primary DENV2 infection in 85 children from the Pediatric Dengue Cohort Study (PDCS) in Managua, Nicaragua. Further, in addition to the gold standard approach to categorize children’s nutritional status, body mass index z-score (BMIz), we explored the use of leptin and adiponectin as blood markers of body fat.

## METHODS

### Ethics statement

The human subjects protocol of the PDCS was reviewed and approved by the institutional review boards of the University of California, Berkeley, and the Nicaraguan Ministry of Health. Parents or legal guardians of all participants provided written informed consent, and participants aged ≥6 years provided assent.

### Study population

The PDCS is an ongoing open cohort study initiated in 2004; every year in March-April, demographic data, weight and height, and healthy non-fasting blood samples are collected from all participants [18]. Blood samples are examined for DENV seroconversion or changes in pre-existing DENV antibody titers using the DENV inhibition enzyme-linked immunosorbent assay (iELISA) as previously described [4,19]. Clinically inapparent DENV infections are identified by seroconversion or a ≥4-fold increase in iELISA titer in 2 consecutive annual samples [4,19]. Cases of clinically apparent DENV infections (i.e., dengue cases) are captured via enhanced passive surveillance and confirmed by reverse transcription-polymerase chain reaction [18,20,21] and/or viral isolation in acute-phase samples or seroconversion of DENV IgM antibodies [18,22] or seroconversion or a ≥4-fold increase in DENV iELISA titers in paired acute- and convalescent-phase samples [4,19]. Children’s nutritional status is anthropometrically defined using BMIz and categorized as normal weight, overweight, or with obesity, in accordance with the World Health Organization Child Growth Standards [16,23]. Among children under 5 years and those aged 5-17 years, normal weight corresponded to BMIz values of -2 to 2 and -2 to 1, respectively; overweight to values above these ranges (>2 or >1); and obesity to BMIz values >3 or >2, respectively [16].

### Study sample

A total of 85 PDCS participants with available pre- and post-infection banked blood samples were included in this study: 31 with normal weight, 30 with overweight, and 24 with obesity. All samples from participants with overweight or obesity who fulfilled our study criteria, experienced a primary DENV2 infection in 2019, and were ≥5 years old were included in the analysis. Participants with normal weight who met study criteria were randomly selected, followed by a stratified selection to ensure a 50–50 representation of clinically inapparent and apparent DENV infections. The peak of DENV2 infections occurred approximately 6±3 months after the 2019 annual sample; thus, pre-infection annual samples (2019) were collected 6±3 months before infection, and post-infection annual samples were collected 6±3 months after infection.

### Laboratory methods

DENV binding antibodies were measured by iELISA, which quantifies circulating total DENV antibodies binding to DENV1-4 antigens and has been described extensively (see Supplemental Material) [4,19]. DENV antigen-specific IgG binding antibodies were measured by a high-throughput multiplex microsphere-based assay (MMBA) that allows measurement of antibodies binding to DENV2 envelope protein (E), envelope domain III (EDIII), and/or non-structural protein 1 (NS1) simultaneously in a single reaction (see Supplemental Material) [24]. The magnitude of IgG antigen-specific binding antibodies was recorded as median fluorescence intensity (MFI) using the iQue3 High-Throughput Screening Cytometry Platform (Sartorius). DENV2 neutralizing antibody titers (NT_50_) were measured using the focus reduction neutralization test (FRNT) assay with mature virions (see Supplemental Material) [25]. The neutralizing efficiency index, i.e., proportion of neutralizing antibodies within binding antibodies, was calculated by dividing the value of neutralizing antibodies by the value of anti-DENV2 E binding IgG antibodies. Leptin and adiponectin were measured in banked annual blood samples collected in 2019, 6±3 months before the infection, using the ELH-Leptin and ELH-Adiponectin ELISA kits (RayBio) following the manufacturer’s recommended protocol.

### Statistical analysis

Statistical models, figures, and tables were generated using R Studio [26]. Correlation analyses between individual host factors (predictors) and log_2_-transformed antibody magnitude (outcome) were performed using linear models. The resulting regression coefficients (β) represent changes in the log_2_-transformed antibody levels per unit increase in the predictor and are reported as fold-changes (2^β^) relative to the reference category for categorical predictors or per unit change for numerical predictors. The overall association between binding and neutralizing antibodies was estimated using linear models. In this model, β represents changes in neutralizing antibody titer per unit increase in binding antibody MFI. Strata-specific β for the three nutritional status categories, three leptin categories, or three adiponectin categories were estimated using multivariate models with an interaction term for the specific type of marker (nutritional status, leptin, or adiponectin). Leptin and adiponectin were categorized into terciles (low, medium, or high) based on participants’ leptin or adiponectin concentration in relation to the distribution of the samples’ leptin or adiponectin values. Percentile-based categories were as follows: low (0-33 percentile), medium-low (33-66 percentile), and high (66-100 percentile). Confidence intervals (CI) were constructed using the Wald method with a critical value of 1.96, producing two-sided 95% CI with a nominal coverage probability of 95%. Two-sided p-values are reported for each coefficient. Given the differences in the ranges of leptin (0.1-8.8 ng/mL) and adiponectin (4.9-80.5 μg/mL) values, coefficient and fold-change estimates are reported per 1-unit increase in leptin and per 10-unit increase in adiponectin. For the interaction linear model, the group-specific coefficients were estimated using the “emtrends” function from the “emmeans” package [27]. Figures and tables displaying model estimates were generated using the “ggplot”, “ggpattern”, “flextable”, and “officer” packages [28–30].

## RESULTS

This study evaluated the association between nutritional status and the antibody response to DENV in 85 children who experienced a primary DENV2 infection in 2019: 31 with normal weight, 30 with overweight, and 24 with obesity (Table 1). Age, sex, and the proportion of clinically apparent infection was balanced between the three nutritional status categories (Table 1). Children with normal weight had the lowest values of circulating leptin, followed by overweight, and then obesity with the highest values (Table 1). Adiponectin was highest in participants with normal weight and lowest in those with obesity (Table 1).

**Table 1.** Participants characteristics.

| Characteristic | Overall | By nutritional status |  |  |
| --- | --- | --- | --- | --- |
|  | n = 85 | Normal weight<br>n = 31 | Overweight<br>n = 30 | Obesity<br>n = 24 |
| **Age<sup>1</sup>** | 9.5 (2.4) | 9.6 (2.5) | 9.7 (2.7) | 9.2 (2.0) |
| **Sex<sup>2</sup>** |  |  |  |  |
| F | 46 (54%) | 15 (48%) | 18 (60%) | 13 (54%) |
| M | 39 (46%) | 16 (52%) | 12 (40%) | 11 (46%) |
| **Infection outcome<sup>2</sup>** |  |  |  |  |
| Inapparent | 47 (55%) | 16 (52%) | 17 (57%) | 14 (58%) |
| Symptomatic | 38 (45%) | 15 (48%) | 13 (43%) | 10 (42%) |
| **Leptin [ng/mL]<sup>1</sup>** | 2.4 (2.3) | 0.8 (0.9) | 2.4 (1.8) | 4.6 (2.3) |
| **Adiponectin [ug/mL]<sup>1</sup>** | 32.7 (13.5) | 35.4 (13.7) | 33.5 (16.4) | 28.4 (7.4) |

<sup>1</sup>Mean (standard deviation)
<sup>2</sup>Count (%)

### Obesity is associated with higher quantities of DENV binding antibodies and lower neutralizing antibody efficiency

The association of nutritional status with antibodies responses was determined using linear regression models adjusted for age, sex, and infection outcome. Children with overweight or obesity had 2.19 times (95%CI 1.22, 3.82) or 1.89 times (95%CI 1.02, 3.48) higher DENV iELISA titers, respectively, compared to participants with normal weight (Figure 1A, Table S1). Obesity was also significantly associated with 1.58 times (95%CI 1.11, 2.26) or 1.65 times (95%CI 1.01, 2.68) higher IgG antigen-specific binding antibodies to EDIII or NS1, respectively (Figure 1A, Table S1). These results were confirmed with a larger sample size, using data from all PDCS participants who experienced a primary or a secondary DENV infection from 2011 to 2022 (688 primary and 937 secondary infections, totaling 1,625 infections) and had available iELISA and BMIz data. With the larger sample size, obesity was again associated with 1.86 times (95%CI 1.20, 2.89; Table S2) higher iELISA antibodies post-primary DENV infection. In children with secondary infections, although those with obesity trended toward higher binding antibodies, the association was not significant in the multivariable analysis (Table S2).

**Figure 1.**
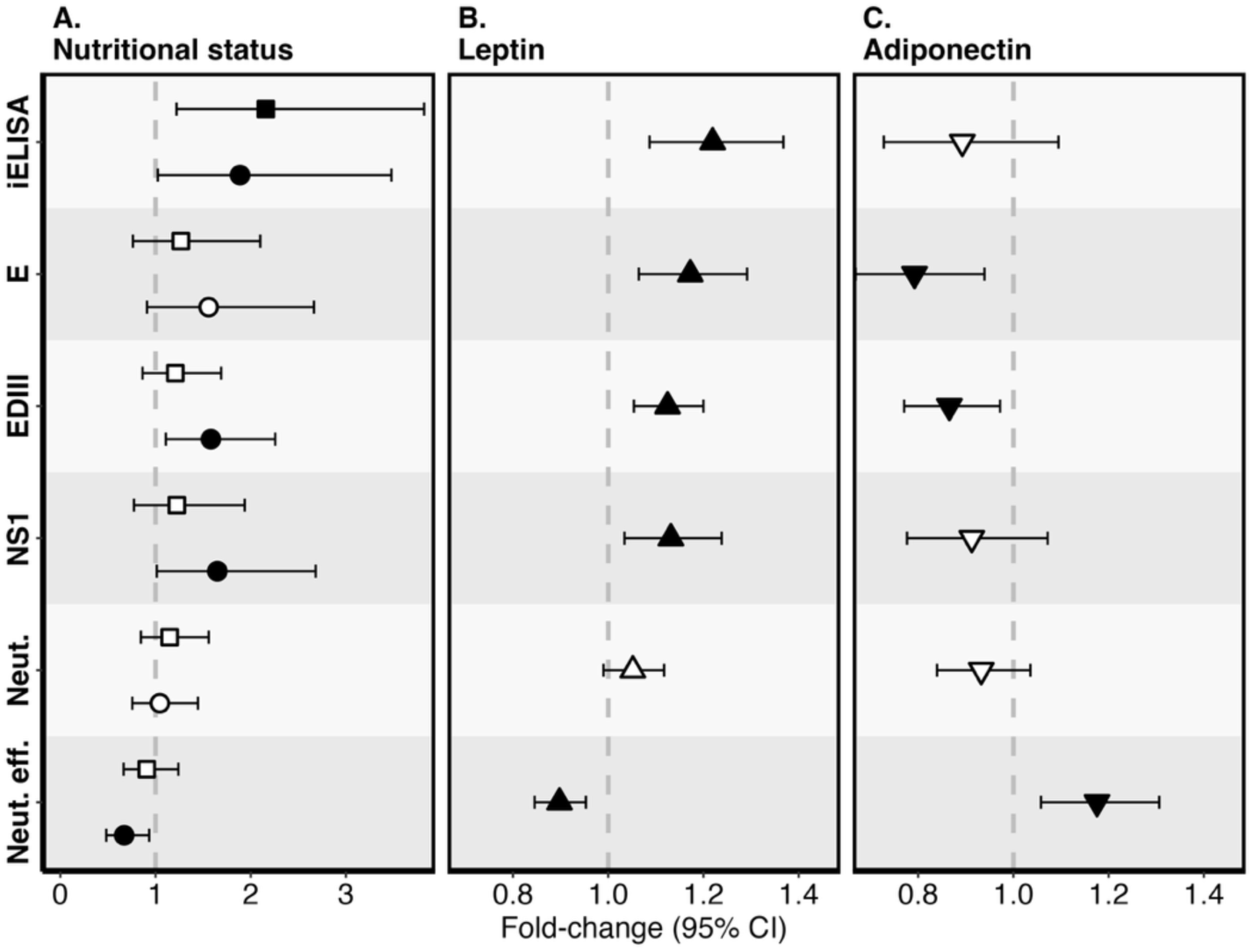
Association between nutritional status and DENV antibody responses. Associations of (**A**) nutritional status, (**B**) leptin, and (**C**) adiponectin with log_2_-transformed antibody responses, estimated using multivariable linear regression models adjusted for age, sex, and infection outcome. Antibody responses included DENV-binding antibodies measured by the inhibition enzyme-linked immunosorbent assay (iELISA); IgG antigen-specific DENV2-binding antibodies to the envelope protein (E), envelope domain III (EDIII), and non-structural protein 1 (NS1), measured by multiplex microsphere-based assay; DENV2 neutralizing antibodies (Neut.) measured by focus reduction neutralization assay; and DENV2 neutralizing efficiency index (Neut. eff.), calculated by dividing the value of neutralizing antibodies by the value of anti-DENV2 E IgG binding antibodies. Regression coefficients (β) were exponentiated (2^β^) and reported as fold-change. Categorical predictors (nutritional status category) are interpreted relative to the reference group (i.e., normal weight), and continuous predictors (i.e., leptin and adiponectin) represent fold-change per unit increase. The dashed grey vertical line indicates the null. Square symbols indicate fold-change for the overweight category; circular symbols represent fold-change for the obesity category; triangle symbols indicate fold-change per unit (ng/mL) increase in leptin; upside-down triangle symbols represent fold-change per 10 unit (μg/mL) increase in adiponectin. Error bars represent 95% confidence intervals. Filled symbols indicate statistically significant predictors (p < 0.05). Open symbols indicate non-significant predictors.

Interestingly, even though children with obesity had higher quantities of binding antibodies, neither overweight nor obesity was associated with neutralizing antibody titers (Figure 1A, Table S1). This discrepancy between higher quantities of binding antibodies but similar levels of neutralizing antibodies led to 0.67 times (95% CI 0.48, 0.93) lower neutralizing efficiency, defined here as the proportion of neutralizing antibodies within E binding antibodies in children with obesity compared to normal weight (Figure 1A, Table S1).

### Higher leptin and lower adiponectin concentration are correlated with higher binding antibodies but not higher neutralizing antibodies

Next, we explored other markers of body fat, such as circulating leptin and adiponectin. Similar to the nutritional status model, a unit increase in leptin concentration (ng/mL) was associated with higher DENV-binding iELISA antibodies (fold-change [FC] 1.22, 95%CI 1.09, 1.37), as well as higher antigen-specific IgG binding antibodies to E (FC 1.17, 95%CI 1.06, 1.29), EDIII (FC 1.13, 95%CI 1.05, 1.20), and NS1 (FC 1.13, 95%CI 1.03, 1.24), but not with neutralizing antibodies (FC 1.05, 95%CI 0.99, 1.12) (Figure 1B, Table S1). Consequently, increases in leptin concentration were associated with 0.90-fold (95%CI 0.85, 0.95) lower neutralization efficiency (Figure 1B, Table S1). A 10-unit increase in adiponectin (ug/mL), which in contrast to leptin is linked to lower body fat [15], was associated with lower binding antibodies to E (FC 0.79, 95%CI 0.67, 0.94) and EDIII FC 0.87, 95%CI 0.77, 0.97) (Figure 1C, Table S1). As with leptin, adiponectin levels were not associated with neutralizing antibodies (FC 0.93, 95%CI 0.84, 1.04) but were associated with higher neutralization efficiency (FC 1.18, 95%CI 1.06, 1.31) (Figure 1B, Table S1).

### Children with obesity need more binding antibodies to reach the same level of neutralizing antibodies

After observing that higher body fat, estimated here by all three surrogate markers (BMIz, leptin, and adiponectin), was associated with lower DENV neutralization efficiency, we modeled the overall and the nutritional status-stratified correlation between binding and neutralizing antibodies. Overall, DENV E-binding IgG antibodies were significantly correlated with DENV neutralizing antibodies (regression coefficient [β] 3.33, 95%CI 2.74, 3.92; Table S3). Interestingly, differences in the regression coefficient were observed by nutritional status category. Children with normal weight showed a significantly (p = 0.02) higher β (4.72, 95%CI 3.54, 5.9) than children with obesity (β 2.68, 95%CI 1.91, 3.44) (Figure 2A-B, Table S3), indicating that children with obesity generate fewer neutralizing antibodies per unit increase in binding antibodies compared to children with normal weight. Similarly, children with the lowest level of leptin showed a significantly (p=0.01) higher β (4.91, 95%CI 3.71, 6.11) when compared to those with the highest level of leptin (β 2.66, 95%CI 1.85, 3.46) (Figure 2C-D, Table S3). Although not statistically significant, this trend aligned with the BMIz and leptin model: those with the highest adiponectin concentrations had a higher β (β 4.75, 95%CI 3.42, 6.08) as compared to those with the lowest concentrations of adiponectin (3.02, 95%CI 2.06, 3.97) (Figure 2E-F, Table S3). This indicates that higher body fat, estimated by multiple proxies—BMIz, higher leptin or lower adiponectin concentrations—is associated with reduced gains in neutralizing antibodies per unit increase of binding antibodies across all models.

**Figure 2.**
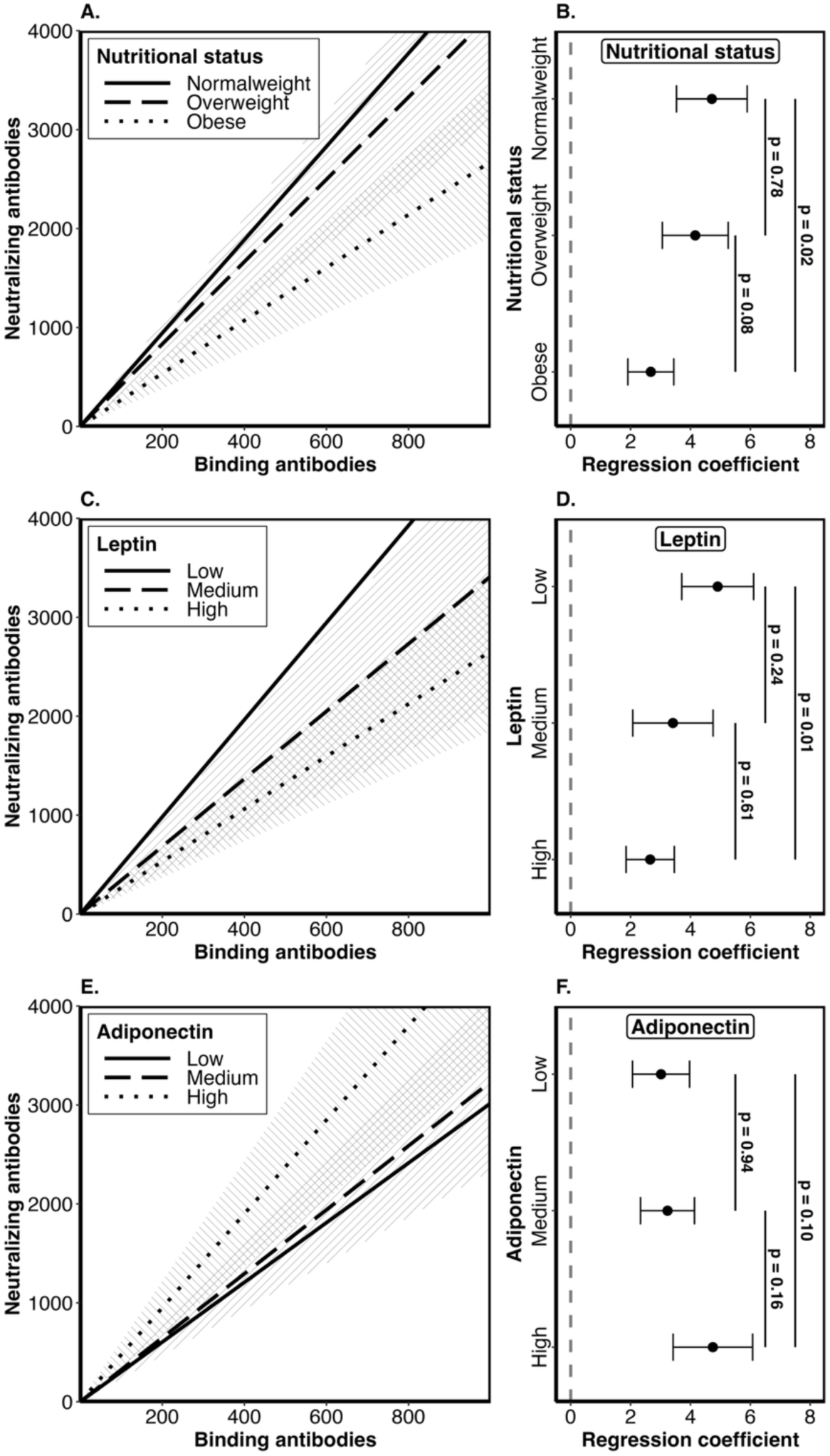
Correlation between binding antibodies and neutralizing antibodies. The correlation between DENV2 envelope protein-binding IgG antibodies and neutralizing antibodies was evaluated using a linear model with an interaction term for (**A-B**) nutritional status category, (**C-D**) leptin category, or (**E-F**) adiponectin category. nutritional status categories were as follows: normal weight, overweight, and obesity. Participants’ leptin or adiponectin concentrations were categorized into terciles: low (0-33 percentile), medium (33-66 percentile), and high (66-100 percentile). Panels A, C, and E show the correlation trend (lines) with 95% confidence intervals (95%CI, shadow) between binding antibodies and neutralizing antibodies, by (**A**) nutritional status category, (**C**) leptin category, or (**E**) adiponectin category. The solid line shows the trend for participants with normal weight, low leptin, or low adiponectin. The dashed lines show the trend for participants with overweight, medium leptin, or medium adiponectin. The dotted lines show the trend for participants with obesity, high leptin, or high adiponectin. In panels B, D, and F, regression coefficients and 95% CI indicate the unit change in neutralizing antibodies per unit change in binding antibodies for each specific category. The dashed grey vertical line indicates the null.

## DISCUSSION

In this study, we report how obesity affects children’s antibody responses after a primary DENV infection using pre- and post-infection demographic data, weight and height measurements, and biological samples from a longitudinal cohort study. Analyses were confirmed using multiple orthogonal approaches. In addition to the BMIz score category typically used to classify children’s nutritional status as normal weight, overweight, and obese, we explored whether two additional proxies of body fat—circulating leptin, which positively correlates with higher body fat, and adiponectin, which negatively correlates with higher body fat—were associated with DENV antibody responses [12,13,15]. Our results indicate that higher body fat, measured by BMIz, leptin, or adiponectin levels, is associated with higher binding antibody levels but not neutralizing antibodies, leading to reduced neutralizing efficiency of the antibody response.

Following primary DENV2 infection, children with obesity produced higher quantities of DENV binding antibodies when compared to children with normal weight. Our results were confirmed using multiple assays—DENV iELISA and DENV MMBA. The iELISA detects total anti-DENV antibodies, without discriminating the antigenic target. Conversely, our MMBA quantifies antigen-specific IgG antibodies binding to DENV2 E, EDIII, or NS1. Consistent findings across iELISA and MMBA suggest that children with obesity have higher DENV-binding antibody responses regardless of antigenic targets examined in this study.

Notably, although children with obesity had a higher quantity of binding antibodies, they did not differ from those with normal weight in terms of neutralizing antibodies. This discrepancy suggests that the antibody response of children with obesity has lower neutralization efficiency; thus, a greater quantity of binding antibodies is needed to reach a similar level of neutralization. This was supported by examining the correlation of binding antibodies with neutralizing antibodies. When compared to children with normal weight, those with obesity had a smaller increase in neutralizing antibodies per unit increase in binding antibodies. The discrepancy between higher binding antibodies but lower neutralization efficiency in children with obesity could be due to differences in the epitopes targeted. DENV virions contain 180 E copies organized in rafts of three E antiparallel homodimers, forming DENV quaternary structure-dependent epitopes [31]. Thus, the quantity of antibodies targeting quaternary epitopes, such as the human monoclonal antibodies 2D22, 1L12, and 3F9, which bind to the major antigenic site of DENV2 neutralizing antibodies, might correlate better with neutralizing antibodies [32,33]. Further, the lower neutralizing potency could be due to generating antibodies with lower affinity. High leptin concentration, as observed in people with obesity, has been shown to reduce transcription of the activation-induced cytidine deaminase (AID) gene [14]. AID is involved in B cell somatic hypermutation; thus, a reduction in AID could lead to lower antibody binding affinity and neutralizing efficiency.

Further, we previously showed that even after accounting for antibody quantity, children with obesity were more likely to be infected with DENV, and once infected, were more likely to develop symptoms, suggesting a lower protective response by DENV antibodies generated by children with obesity [7]. People with greater body surface are more likely to be bitten by mosquitoes [34,35] and have been associated with higher ZIKV seropositivity [36]. Such increased mosquito biting could lead to a higher DENV inoculum, providing a greater antigen load—thus leading to higher B cell activation frequency and subsequently to a greater quantity of antibodies. Additionally, children with obesity are at higher risk of developing dengue disease and progressing to severe disease [7–9]. Therefore, clinical or subclinical differences in the course of infection, such as higher viral load or greater inflammatory responses in children with obesity, could influence the antibody response.

A notable strength of this study was the use of multiple approaches to estimate the effect of nutritional status on anti-DENV antibody responses, all showing a similar association with the antibody response. In addition to the gold standard approach for nutritional status, BMIz categorization, we validated our results using two known blood proxies of body fat. Leptin and adiponectin were chosen because of their relationship with body fat and their consistently different concentrations between individuals with normal weight and obesity, despite postprandial fluctuations [15,37]. Supporting our analysis with the BMIz method, higher leptin was associated with higher binding antibodies and with lower neutralization efficacy. Markedly, adiponectin displayed a pattern similar to leptin association with the antibody responses, but in the inverse direction. Increases in adiponectin were associated with lower antibody quantity but with higher quality. Given that circulating leptin concentration increases with higher body fat while adiponectin levels decrease, the contrary direction in the estimated coefficient between the leptin and adiponectin model is expected and further validates our results. Further, the prospective cohort study design of the PDCS, which collects data and banks blood samples annually, allowed us to measure exposures of interest, such as BMIz, leptin, and adiponectin, before the studied outcome occurred. This approach respects the vital principle that a cause must precede its effect. Moreover, our analysis differentiated antibody quantity (i.e., binding) from antibody quality (i.e., neutralization). This allowed us to identify a distinct association pattern between nutritional status and the quantity or quality of antibodies. Lastly, we confirmed our results between obesity and higher DENV iELISA binding antibodies using data from 1,625 children from the PDCS, including both primary and secondary infections.

Nonetheless, this study has several limitations. The MMBA measures antibodies binding to the monomeric form of E and cannot discriminate antibodies binding to specific antigenic sites within the E protein, such as DENV quaternary epitopes present only on E dimers or the virion. Further, given that this is an observational study with natural DENV infections and since people with larger bodies are more likely to be bitten by mosquitoes, we cannot distinguish whether the higher quantity of binding antibodies observed here in children with obesity is caused directly by higher body fat or indirectly by a higher risk of mosquito bites in children with obesity. Randomized controlled trials, most likely clinical vaccine trials, where all participants receive the same inoculum of DENV antigen, could directly answer this question by including participants with distinct nutritional statuses. Moreover, given the prospective study design, where the time of infections cannot be controlled, our time points pre- and post-infection are imprecise. The peak of the infection happened 6 months after the pre-infection sample and 6 months before the post-infection sample was collected; however, sampling time points may vary by approximately ±3 months from the indicated interval, increasing heterogeneity (i.e., 95%CI) in our association results.

In conclusion, here we show that nutritional status, estimated using multiple proxies, affects antibody responses post-primary DENV2 infection. Higher body fat, in the form of obesity, high leptin, or low adiponectin concentrations, is associated with a higher quantity of DENV-binding antibodies but not with higher neutralizing antibodies. This suggests that antibodies generated by children with obesity have a lower neutralizing efficiency, and a higher quantity of binding antibodies is needed to reach the same quality observed in those with normal weight. Given that non-neutralizing DENV-binding antibodies have the potential to enhance risk of disease in a subsequent DENV infection via ADE, and that obesity, even after adjusting for pre-existing antibodies, has also been linked to a higher risk of dengue and severe dengue, it is important to further explore these associations [4–9]. Collectively, leptin and adiponectin demonstrated consistent associations with DENV antibody responses mirroring those observed with BMIz, thus establishing these biomarkers as viable blood-based proxies of body fat for immunological studies. This is particularly valuable when height and weight measurements are unavailable and antibody responses, along with leptin and adiponectin, can be measured in banked samples. In the absence of BMIz measurements, leptin and adiponectin can be measured in banked blood samples and used as proxies of body fat. This and other studies have shown the importance of considering nutritional status in infectious disease transmission, pathogenesis, and immunology research.

## Notes

### Authors Contributions

R.M.H. and E.H. conceptualized the study. A.B. and G.K. supervised the cohort study. R.M.H. and S.B. developed the multiplex microsphere-based assay. R.M.H. generated the data, conducted the statistical and created the figures and tables. R.M.H. wrote the initial manuscript draft. E.H. edited the manuscript. S.B., A.B., and G.K. reviewed the article for intellectual content.

## Acknowledgments

We thank the personnel at the Sustainable Sciences Institute, the Laboratorio Nacional de Virología, and the Centro de Salud Sócrates Flores Vivas in Managua, Nicaragua, for their excellent work and dedication to the PDCS. We are grateful to all the children in the PDCS and their families, without whom this study would not have been possible. We are thankful to Dr. Lakshmanane Premkumar at the University of North Carolina at Chapel Hill for his generous donation of DENV EDIII protein. We thank Dr. Nina Holland at the University of California Berkeley for her advice regarding the leptin and adiponectin assays.

## Data availability

After securing approval from the University of California–Berkeley Committee for the Protection of Human Subjects (CPHS), individual data for figure reproduction can be shared with external researchers. For data access arrangements, please contact E.H. at or the CPHS at. Standard data and material transfer agreements govern all materials and data used in this study.

## SUPPLEMENTARY MATERIAL

## SUPPLEMENTARY METHODS

### Inhibition ELISA to measure DENV binding antibodies

The inhibition ELISA (iELISA) assay, which quantifies total circulating DENV antibodies binding to DENV serotype 1-4 antigens, has been extensively described [1,2]. Briefly, upon enrollment and in every annual sample thereafter, DENV antibodies are measured for each PDCS participant using the iELISA at a single dilution (1 in 10). If DENV antibodies are detected (i.e., positive iELISA signal at 1/10), the iELISA titer (see below) is estimated by testing serial 10-fold dilutions of the samples. Once a participant has a positive iELISA result, iELISA titers are determined for the current and all subsequent annual samples, and results are reported as the reciprocal dilution factor that inhibits 50% of the binding of a standardized polyclonal mix of DENV-positive samples. Since iELISA results are log_2_-transformed for modeling, negative raw iELISA results (i.e., no signal at 1/10 dilution) are reported as a titer of 2. Positive iELISA titers are reported as the geometric mean of two technical replicate titers, one performed in the current year and the second conducted the following year. The raw iELISA data were generated and provided by the PDCS team in Managua, Nicaragua. A detailed iELISA protocol can be found in Katzelnick et al. [1].

### Multiplex microsphere-based assay for antigen-specific antibodies

The DENV multiplex Luminex-based assay is a high-throughput multiplex microsphere-based assay (MMBA) that enables the measurement of IgG antigen-specific antibodies binding to DENV envelope protein (E), envelope domain III (EDIII), and non-structural protein 1 (NS1) simultaneously in a single reaction and has been previously described [3]. Here only DENV2 antigens were analyzed. The E and NS1 proteins were sourced from The Native Antigen Company. Briefly, both E and NS1 were biotinylated using the EZ-Link Sulfo NHS-LC-LC-Biotin (Thermo Fisher Scientific) at 180.5 ng of biotin per ug of protein. EDIII, with a single site-specific biotinylation, was provided by Lakshmanane Premkumar (University of North Carolina at Chapel Hill) [4]. Biotinylated antigens were conjugated to a specific avidin-coated MagPlex magnetic microsphere region (Luminex Corporation) at 250 ng per 100,000 microspheres. Participants’ serum samples were diluted 1/800 in PBS with 1% BSA, then 20uL of diluted serum was mixed with 20uL the multiplex microsphere mix containing 500 microspheres per each region in a 384-well plate (Greiner), incubated for 90 minutes at 1,200 RPM shaking at 37°C in a digital microplate shaker (Thermo Fisher Scientific), and washed three times with 1X PBS, 0.01% BSA, 0.02% Tween-20 (washing buffer) using an automatic plate washer (Tecan Hydrospeed). The magnitude of IgG antigen-specific binding antibodies was determined using phycoerythrin-tagged anti-human secondary antibodies (Southern Biotech). The phycoerythrin signal per microsphere region, representing the level of antibodies binding to the specific protein conjugated to each microsphere region, was recorded as median fluorescence intensity (MFI) using the iQue3 High-Throughput Screening Cytometry Platform (Sartorius).

### Focus reduction neutralization assay for DENV neutralizing antibodies

DENV2 neutralizing antibody titers (NT_50_) were measured using the focus reduction neutralization test (FRNT) assay. Vero cells (ATCC CCL-81), maintained in culture medium (minimum essential medium [Gibco] with 5% fetal bovine serum, 1% nonessential amino acids [Gibco], 1% sodium pyruvate [Gibco], and 1% penicillin-streptomycin [Gibco]), were seeded in a 96-well flat-bottom tissue culture-treated plate at 15,000 cells per well and incubated overnight at 37°C in 5% CO_2_. The DENV inoculum solution was prepared using a clinical isolate from Nicaragua (DENV2, 8891.12a1SPD2) grown in Vero-Furin cells [5], at a concentration of 150 focus-forming units per well. Participants’ sera were heat-inactivated at 56°C for 30 minutes, serially diluted (seven-point three-fold serial dilution from 1/10 to 1/7,290), and incubated with an equal volume of DENV2 solution for 1 hour at 37°C in 5% CO_2_. After removal of the Vero cell culture medium, the cells were incubated in 30 μL of serum-DENV solution for 1.5 hours at 37°C. Then, 40 μL of growth medium was added and incubated for 30 hours at 37°C. Vero cells were fixed in 4% formaldehyde by adding 70 μL of 8% formaldehyde to the 70 μL of cell medium for 15 minutes at room temperature. Cells were washed with 1X PBS, the formaldehyde solution was discarded, and cells were permeabilized with 50 μL 0.5% Triton for 6 minutes at room temperature. Cells were washed twice in washing buffer (1% BSA in PBS). DENV-infected cell foci were stained with 50 uL of mouse anti-DENV envelope 4G2 monoclonal antibody (Biomatik) at 1/1,000 overnight at 4°C, washed twice, and incubated with 50 μL horseradish peroxidase-conjugated goat anti-mouse secondary antibody (Biolegend) diluted at 1/2,000 for 1 hour at room temperature. Cells were washed twice with PBS, incubated with 50 μL KLP TrueBlue Peroxidase Substrate (Seracare) for 30 minutes at room temperature, and washed with tap water. DENV-infected cell foci were counted using a CTL Immunospot analyzer and the BioSpot software (Cellular Technology, Ltd.). Using the foci count from the serially diluted samples, participants’ 50% neutralization titers (NT_50_), the reciprocal of the plasma dilution that achieved a 50% reduction in infection, were generated by fitting a sigmoidal dose-response curve in R Studio using the drda package [6]. The neutralizing efficiency index, i.e., proportion of neutralizing antibodies within the binding antibodies, was calculated by dividing the value of neutralizing antibodies by the value of anti-DENV2 E IgG binding antibodies.

## SUPPLEMENTARY TABLES

**Table S1.** Nutritional status association with the antibody response post-primary DENV infection in the 85 study participants.

| Model | Dependent | Independent | Coefficient (95% CI)<sup>1</sup> | Fold-change (95% CI)<sup>2</sup> | p-value |
| --- | --- | --- | --- | --- | --- |
| Bivariate | iELISA | Overweight | 1.13 (0.321, 1.939) | 2.189 (1.249, 3.834) | 0.008 |
| Bivariate | Envelope | Overweight | 0.426 (-0.315, 1.167) | 1.344 (0.804, 2.246) | 0.263 |
| Bivariate | EDIII | Overweight | 0.343 (-0.153, 0.839) | 1.268 (0.9, 1.789) | 0.179 |
| Bivariate | NS1 | Overweight | 0.349 (-0.31, 1.008) | 1.274 (0.807, 2.011) | 0.302 |
| Bivariate | Neutralization | Overweight | 0.267 (-0.184, 0.717) | 1.203 (0.88, 1.644) | 0.249 |
| Bivariate | Neut. eff. | Overweight | -0.157 (-0.612, 0.298) | 0.897 (0.654, 1.229) | 0.500 |
| Bivariate | iELISA | Obesity | 0.955 (0.09, 1.82) | 1.938 (1.064, 3.531) | 0.034 |
| Bivariate | Envelope | Obesity | 0.651 (-0.136, 1.438) | 1.571 (0.91, 2.71) | 0.109 |
| Bivariate | EDIII | Obesity | 0.678 (0.152, 1.205) | 1.6 (1.111, 2.305) | 0.013 |
| Bivariate | NS1 | Obesity | 0.746 (0.047, 1.446) | 1.678 (1.033, 2.724) | 0.040 |
| Bivariate | Neutralization | Obesity | 0.088 (-0.39, 0.566) | 1.063 (0.763, 1.481) | 0.719 |
| Bivariate | Neut. eff. | Obesity | -0.563 (-1.046, -0.08) | 0.677 (0.484, 0.946) | 0.025 |
| Bivariate | iELISA | Leptin | 0.243 (0.087, 0.398) | 1.183 (1.062, 1.318) | 0.003 |
| Bivariate | Envelope | Leptin | 0.249 (0.12, 0.379) | 1.189 (1.086, 1.3) | 0.000 |
| Bivariate | EDIII | Leptin | 0.185 (0.098, 0.272) | 1.137 (1.071, 1.208) | 0.000 |
| Bivariate | NS1 | Leptin | 0.178 (0.059, 0.298) | 1.132 (1.041, 1.229) | 0.005 |
| Bivariate | Neutralization | Leptin | 0.102 (0.02, 0.184) | 1.073 (1.014, 1.136) | 0.016 |
| Bivariate | Neut. eff. | Leptin | -0.146 (-0.227, -0.064) | 0.904 (0.854, 0.956) | 0.001 |
| Bivariate | iELISA | Adipo | -0.105 (-0.37, 0.16) | 0.93 (0.774, 1.117) | 0.439 |
| Bivariate | Envelope | Adipo | -0.355 (-0.581, -0.128) | 0.782 (0.668, 0.915) | 0.003 |
| Bivariate | EDIII | Adipo | -0.216 (-0.373, -0.06) | 0.861 (0.772, 0.96) | 0.008 |
| Bivariate | NS1 | Adipo | -0.139 (-0.351, 0.073) | 0.908 (0.784, 1.052) | 0.202 |
| Bivariate | Neutralization | Adipo | -0.107 (-0.249, 0.035) | 0.928 (0.841, 1.025) | 0.144 |
| Bivariate | Neut. eff. | Adipo | 0.246 (0.107, 0.385) | 1.186 (1.077, 1.306) | 0.001 |
| Multivariate | iELISA | Overweight | 1.11 (0.287, 1.934) | 2.159 (1.22, 3.821) | 0.010 |
| Multivariate | Envelope | Overweight | 0.34 (-0.391, 1.07) | 1.265 (0.763, 2.099) | 0.365 |
| Multivariate | EDIII | Overweight | 0.272 (-0.212, 0.756) | 1.208 (0.864, 1.689) | 0.274 |
| Multivariate | NS1 | Overweight | 0.292 (-0.37, 0.954) | 1.224 (0.774, 1.937) | 0.390 |
| Multivariate | Neutralization | Overweight | 0.199 (-0.242, 0.64) | 1.148 (0.846, 1.559) | 0.378 |
| Multivariate | Neut. eff. | Overweight | -0.139 (-0.588, 0.31) | 0.908 (0.665, 1.24) | 0.545 |
| Multivariate | iELISA | Obesity | 0.916 (0.034, 1.798) | 1.887 (1.024, 3.478) | 0.045 |
| Multivariate | Envelope | Obesity | 0.639 (-0.135, 1.414) | 1.558 (0.911, 2.664) | 0.110 |
| Multivariate | EDIII | Obesity | 0.661 (0.148, 1.174) | 1.581 (1.108, 2.257) | 0.014 |
| Multivariate | NS1 | Obesity | 0.722 (0.02, 1.423) | 1.649 (1.014, 2.682) | 0.047 |
| Multivariate | Neutralization | Obesity | 0.064 (-0.404, 0.531) | 1.045 (0.756, 1.445) | 0.791 |
| Multivariate | Neut. eff. | Obesity | -0.576 (-1.052, -0.1) | 0.671 (0.482, 0.933) | 0.020 |
| Multivariate | iELISA | Leptin | 0.286 (0.12, 0.452) | 1.219 (1.087, 1.368) | 0.001 |
| Multivariate | Envelope | Leptin | 0.229 (0.09, 0.369) | 1.172 (1.064, 1.292) | 0.002 |
| Multivariate | EDIII | Leptin | 0.169 (0.076, 0.263) | 1.125 (1.054, 1.2) | 0.001 |
| Multivariate | NS1 | Leptin | 0.178 (0.048, 0.308) | 1.132 (1.034, 1.238) | 0.009 |
| Multivariate | Neutralization | Leptin | 0.073 (-0.014, 0.16) | 1.052 (0.99, 1.117) | 0.106 |
| Multivariate | Neut. eff. | Leptin | -0.156 (-0.242, -0.069) | 0.898 (0.845, 0.953) | 0.001 |
| Multivariate | iELISA | Adipo | -0.164 (-0.458, 0.13) | 0.893 (0.728, 1.095) | 0.278 |
| Multivariate | Envelope | Adipo | -0.336 (-0.581, -0.091) | 0.792 (0.668, 0.939) | 0.009 |
| Multivariate | EDIII | Adipo | -0.208 (-0.376, -0.041) | 0.865 (0.771, 0.972) | 0.017 |
| Multivariate | NS1 | Adipo | -0.132 (-0.364, 0.1) | 0.913 (0.777, 1.072) | 0.269 |
| Multivariate | Neutralization | Adipo | -0.101 (-0.252, 0.051) | 0.933 (0.84, 1.036) | 0.196 |
| Multivariate | Neut. eff. | Adipo | 0.233 (0.081, 0.386) | 1.176 (1.058, 1.306) | 0.004 |

Abbreviations: 95% CI, 95% confidence interval; iELISA; DENV binding antibodies measured by inhibition ELISA; E; DENV2 envelope protein binding antibodies measured by Luminex; EDIII, DENV2 envelope protein domain III binding antibodies measured by Luminex; NS1, DENV2 non-structural protein 1 binding antibodies measured by Luminex; Neutralization; DENV2 neutralizing antibodies measured by FRNT assay; Neut. eff.; DENV2 neutralizing efficiency (neutralizing antibodies divided by E-binding antibodies).
<sup>1</sup>Estimated coefficient in log<sub>2</sub> scale; represents exponential changes with base 2 in the dependent variable in relation to the independent variable reference value for categorical predictor or by unit change for numerical predictor.
<sup>2</sup>Fold-change (2<sup>estimated coefficient</sup>); represents fold-changes in the dependent variable in relation to the independent variable reference value for categorical predictor or by unit change for numerical predictor.

**Table S2.**
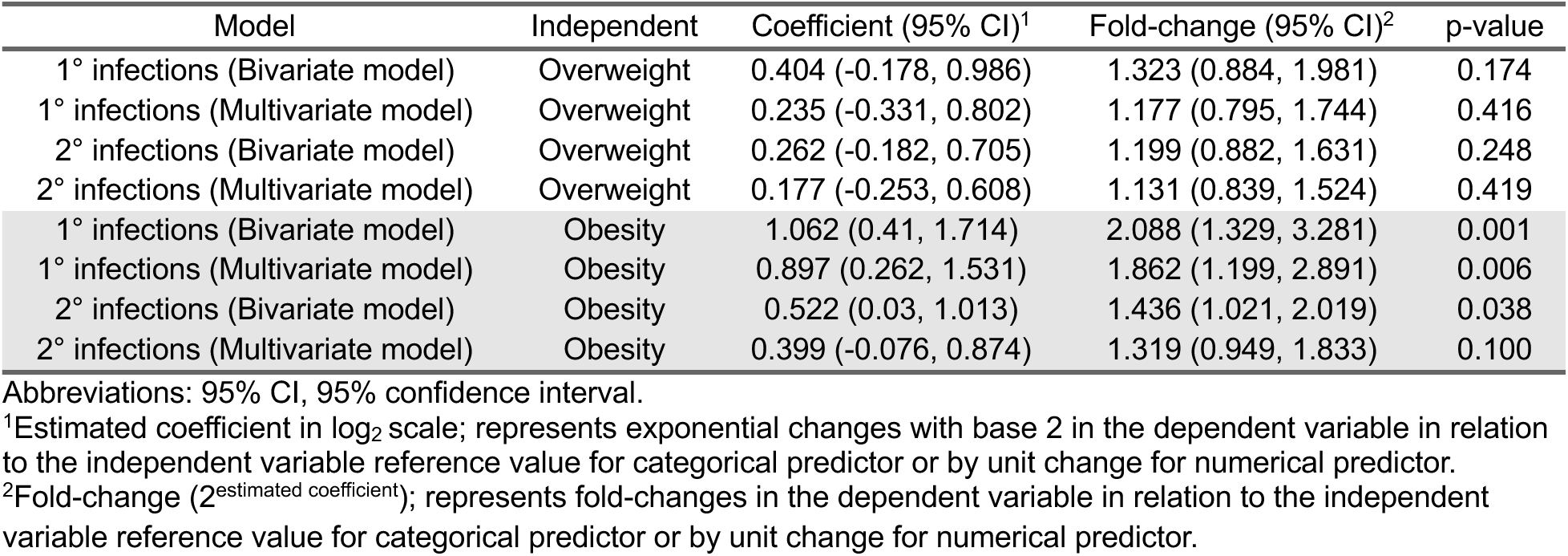
Nutritional status association with the antibody response post-primary or post-secondary DENV infection using all available data from the PDCS.

**Table S3.** Correlation between binding and neutralizing antibodies.

| Interaction | Independent | Coefficient (95% CI)<sup>1</sup> | P value |
| --- | --- | --- | --- |
| BMIz | Overall | 3.332 (2.744, 3.92) | 4.37e-18 |
| BMIz | Normalweight | 4.715 (3.535, 5.896) | 1.88e-11 |
| BMIz | Overweight | 4.161 (3.061, 5.261) | 1.20e-10 |
| BMIz | Obese | 2.676 (1.909, 3.443) | 1.50e-09 |
| Leptin | Low | 4.91 (3.709, 6.111) | 8.22e-12 |
| Leptin | Medium | 3.416 (2.074, 4.757) | 3.53e-06 |
| Leptin | High | 2.655 (1.85, 3.461) | 7.85e-09 |
| Adiponectin | Low | 3.017 (2.064, 3.971) | 2.48e-08 |
| Adiponectin | Medium | 3.234 (2.333, 4.135) | 6.78e-10 |
| Adiponectin | High | 4.748 (3.421, 6.076) | 7.54e-10 |

Abbreviations: 95% CI, 95% confidence interval.
<sup>1</sup>Estimated coefficient represents a unit change in DENV2 neutralizing antibodies per unit change in DENV2 E-specific IgG binding antibodies.

## References

1. Cattarino L, Rodriguez-Barraquer I, Imai N et al. Mapping global variation in dengue transmission intensity. Sci Transl Med. 2020;12:eaax4144.

2. World Health Organization, editor. Dengue haemorrhagic fever: diagnosis, treatment, prevention, and control. 2nd ed. Geneva: World Health Organization; 1997.

3. Special Programme for Research and Training in Tropical Diseases, World Health Organization, editors. Dengue: guidelines for diagnosis, treatment, prevention, and control. New ed. Geneva: TDR: World Health Organization; 2009.

4. Katzelnick LC, Gresh L, Halloran ME et al. Antibody-dependent enhancement of severe dengue disease in humans. Science. 2017; 358:929–32.

5. Katzelnick LC, Narvaez C, Arguello S et al. Zika virus infection enhances future risk of severe dengue disease. Science. 2020; 369:1123–8.

6. Waggoner JJ, Katzelnick LC, Burger-Calderon R et al. Antibody-Dependent Enhancement of Severe Disease Is Mediated by Serum Viral Load in Pediatric Dengue Virus Infections. The Journal of Infectious Diseases. 2020; 221:1846–54.

7. Mercado-Hernandez R, Myers R, Bustos Carillo F et al. Obesity Is Associated With Increased Pediatric Dengue Virus Infection and Disease: A 9-Year Cohort Study in Managua, Nicaragua. Clin. Infect. Dis. 2024; 79:1102–1108.

8. Zulkipli MS, Dahlui M, Jamil N et al. The association between obesity and dengue severity among pediatric patients: A systematic review and meta-analysis. PLoS Negl Trop Dis. 2018; 12:e0006263.

9. Chen CY, Chiu YY, Chen YC et al. Obesity as a clinical predictor for severe manifestation of dengue: a systematic review and meta-analysis. BMC Infect Dis. 2023; 23:502.

10. Nieman, D, Henson D, Nehlsen-Cannarella S et al. Influence of obesity on immune function. J Am Diet Assoc. 1999; 99:294–9.

11. Lumeng CN, Bodzin JL, Saltiel AR. Obesity induces a phenotypic switch in adipose tissue macrophage polarization. J Clin Invest. 2007; 117:175–84.

12. Considine RV, Kriauciunas A, Ohannesian JP. Serum Immunoreactive-Leptin Concentrations in Normal-Weight and Obese Humans. N Engl J Med. 1996; 334:4.

13. Dencker M, Thorsson O, Karlsson M, Lindén C, Wollmer P, Ahrén B. Leptin is closely related to body fat in prepubertal children aged 8–11 years. Acta Paediatrica. 2006; 95:975–9.

14. Frasca D, Diaz A, Romero M, Blomberg BB. Leptin induces immunosenescence in human B cells. Cell. Immunol. 2020; 348:103994.

15. Rocha ARDF, Morais NDSD, Azevedo FM et al. Leptin, CRP, and adiponectin correlate with body fat percentage in adolescents: systematic review and meta-analysis. Front Nutr. 2025; 12:1560080.

16. WHO Obesity and overweight key facts. WHO Obesity and overweight key facts. 2025. WHO Obesity and overweight key facts.

17. NCD Risk Factor Collaboration (NCD-RisC). Worldwide trends in body-mass index, underweight, overweight, and obesity from 1975 to 2016: a pooled analysis of 2416 population-based measurement studies in 128·9 million children, adolescents, and adults. The Lancet. 2017; 390:2627–42.

18. Kuan G, Gordon A, Avilés W et al. The Nicaraguan pediatric dengue cohort study: study design, methods, use of information technology, and extension to other infectious diseases. Am J Epidemiol. 2009; 170:120–9.

19. Balmaseda A, Hammond SN, Tellez Y et al. High seroprevalence of antibodies against dengue virus in a prospective study of schoolchildren in Managua, Nicaragua. Trop Med Int Health. 2006; 11:935–42.

20. Waggoner JJ, Abeynayake J, Sahoo MK et al. Single-reaction, multiplex, real-time rt-PCR for the detection, quantitation, and serotyping of dengue viruses. PLoS Negl Trop Dis. 2013; 7:e2116.

21. Waggoner JJ, Gresh L, Mohamed-Hadley A et al. Single-Reaction Multiplex Reverse Transcription PCR for Detection of Zika, Chikungunya, and Dengue Viruses. Emerg Infect Dis. 2016; 22:1295– 7.

22. Balmaseda A, Guzmán MG, Hammond S et al. Diagnosis of Dengue Virus Infection by Detection of Specific Immunoglobulin M (IgM) and IgA Antibodies in Serum and Saliva. Clin Diagn Lab Immunol. 2003; 10:317–22.

23. WHO Multicentre Growth Reference Study Group. Length/height-for-age, weight-for-age, weight-for-length, weight-for-height and body mass index-for-age; methods and development. Onis M de, editor. Geneva: WHO Press; 2006.

24. Bos S, Singh T, Zambrana JV et al. Longitudinal antibody profiling after dengue reveals distinct dynamics by antibody specificity. Nat Commun. In press.

25. Bos S, Graber AL, Cardona-Ospina JA et al. Protection against symptomatic dengue infection by neutralizing antibodies varies by infection history and infecting serotype. Nat Commun. 2024; 15:382.

26. R Core Team. 2023. R: A Language and Environment for Statistical Computing. R Foundation for Statistical Computing, Vienna, Austria. Version 4.3.1.

27. Lenth RV. emmeans: Estimated Marginal Means, aka Least-Squares Means. 2017. Available from: https://CRAN.R-project.org/package=emmeans doi:10.32614/CRAN.package.emmeans

28. Fc M, Davis TL, ggplot2 authors. ggpattern: “ggplot2” Pattern Geoms. 2022. Available from: https://CRAN.R-project.org/package=ggpattern doi:10.32614/CRAN.package.ggpattern

29. Gohel D, Skintzos P. flextable: Functions for Tabular Reporting. 2017. Available from: https://CRAN.R-project.org/package=flextable doi:10.32614/CRAN.package.flextable

30. Gohel D, Moog S, Heckmann M. Officer: Manipulation of Microsoft Word and PowerPoint Documents. 2017. Available from: https://CRAN.R-project.org/package=officer doi:10.32614/CRAN.package.officer

31. Wahala WMPB, De Silva AM. The Human Antibody Response to Dengue Virus Infection. Viruses. 2011; 3:2374–95.

32. Gallichotte EN, Baric TJ, Yount BL et al. Human dengue virus serotype 2 neutralizing antibodies target two distinct quaternary epitopes. Barrett ADT, editor. PLoS Pathog. 2018; 14:e1006934.

33. Gallichotte EN, Widman DG, Yount BL et al. A New Quaternary Structure Epitope on Dengue Virus Serotype 2 Is the Target of Durable Type-Specific Neutralizing Antibodies. mBio. 2015; 6:e01461–15.

34. Port GR, Boreham PFL, Bryan JH. The relationship of host size to feeding by mosquitoes of the *Anopheles gambiae* Giles complex (Diptera: Culicidae). Bull Entomol Res. 1980; 70:133–44.

35. Harrington LC, Fleisher A, Ruiz-Moreno D et al. Heterogeneous Feeding Patterns of the Dengue Vector, Aedes aegypti, on Individual Human Hosts in Rural Thailand. Bingham A, editor. PLoS Negl Trop Dis. 2014; 8:e3048.

36. Zambrana JV, Bustos Carrillo F, Burger-Calderon R et al. Seroprevalence, risk factor, and spatial analyses of Zika virus infection after the 2016 epidemic in Managua, Nicaragua. Proc Natl Acad Sci USA. 2018; 115:9294–9.

37. Larsen MA, Isaksen VT, Paulssen EJ et al. Postprandial leptin and adiponectin in response to sugar and fat in obese and normal weight individuals. Endocrine. 2019; 66:517–25.

## REFERENCES

1. Katzelnick LC, Gresh L, Halloran ME et al. Antibody-dependent enhancement of severe dengue disease in humans. Science. 2017; 358:929–32.

2. Balmaseda A, Hammond SN, Tellez Y et al. High seroprevalence of antibodies against dengue virus in a prospective study of schoolchildren in Managua, Nicaragua. Trop Med Int Health. 2006; 11:935–42.

3. Bos S, Singh T, Zambrana JV et al. Longitudinal antibody profiling after dengue reveals distinct dynamics by antibody specificity. Nat Commun. In press.

4. Hein LD, Castillo IN, Medina FA et al. Multiplex sample-sparing assay for detecting type-specific antibodies to Zika and dengue viruses: an assay development and validation study. The Lancet Microbe. 2025; 6:100951.

5. Bos S, Graber AL, Cardona-Ospina JA et al. Protection against symptomatic dengue infection by neutralizing antibodies varies by infection history and infecting serotype. Nat Commun. 2024; 15:382.

6. Malyutina A, Tang J, Pessia A. drda : An *R* Package for Dose-Response Data Analysis Using Logistic Functions. J Stat Soft. 2023;106(4).

